# A Novel Approach to Brain Tumor Classification Using Deep Neural Networks

**DOI:** 10.1101/2023.10.03.23296522

**Authors:** Rohan Tummala

## Abstract

The American Cancer Society estimates that in 2023 about 24,810 malignant brain and spinal cord tumors will be diagnosed in the United States and about 18,990 people will die from these tumors. Early detection of malignant brain tumors is crucial in improving survival. Currently, MRI is the standard radiological method used to identify brain tumors. However, these tests are prone for human error, inefficiencies, and often require invasive tissue diagnosis for confirmation. Advancement in machine learning algorithms has shown to improve accuracy in detection of brain tumors. In this study, a novel deep learning approach, called Inception Resnet, a type of pretrained Convolutional Neural Network(CNN), was used to identify and classify three common brain tumors. These classes were pituitary, meningioma, and glioma. Pituitary tumors are the most harmless of the three as they are noncancerous. This contrasts with both glioma and high grade meningioma tumors which are life-threatening. It should be noted, however, that high grade meningioma tumors are very rare. Along with these three classes, a separate control class with no brain tumors was also used. This dataset contained 5,952 MRI images that included 1621 glioma images, 574 meningioma images, 1751 pituitary images and 2000 images with no tumor. Based on this data, a model was created that was able to diagnose brain tumors with an accuracy of 96.7%.

## Introduction

Malignant brain tumors are one of the most lethal forms of cancer. The American Cancer Society estimates that in 2023, in the United States, about 24,810 malignant tumors of the brain and spinal cord will be diagnosed and about 18,990 people will die from these tumors^1^. Early detection of malignant brain tumors is crucial in improving survival. Currently, MRI (Magnetic Resonance Imaging) is the standard radiological method used to identify brain tumors. However, these tests are prone for human error, inefficiencies and most often require invasive tissue diagnosis for confirmation^2^. Advancement in Artificial intelligence and machine learning algorithms has been shown to improve accuracy in the detection of brain tumors. The advantages to using a machine learning model to autonomously detect the presence and type of tumors in patients include speed, efficiency, accuracy, and using fewer resources. The purpose of this study is to use a novel machine learning model, using Inception Resnet, a type of Convolutional Neural Network (CNN) to identify and classify three common types of brain tumors. These classes were pituitary, meningioma, and glioma. Pituitary tumors are the most harmless of the three as they are noncancerous. This contrasts with both glioma and high grade meningioma tumors which are life-threatening^3^. Along with these three classes, a separate control class with no brain tumors was also included in the dataset.

### Dataset

The data was obtained from the open-source Kaggle database. The dataset included MRI images of both healthy and brain tumor patients. The dataset used in this model is a set of 5,952 images of MRI scans taken from a variety of angles. MRI stands for Magnetic Resonance Imaging, and, as the name suggests, implements a large, tube shaped magnet that realigns water molecules in a patient’s body^4^. Radio waves then cause the aligned atoms to produce signals, which are represented on the final image. The specific dataset used contains images that are labeled as belonging to one of four classes. Three of them are brain cancer types—pituitary, meningioma, glioma, and the fourth class has no tumor. In total, there were 1621 glioma images, 574 meningioma images, 1751 pituitary images and 2000 images with no tumor. The images were taken in three different orientations: sagittal, coronal, and transverse. As seen in Figure 1, the more lethal glioma and meningioma tumors are much bigger whereas the pituitary tumor is mostly hidden.

**Figure 1:**
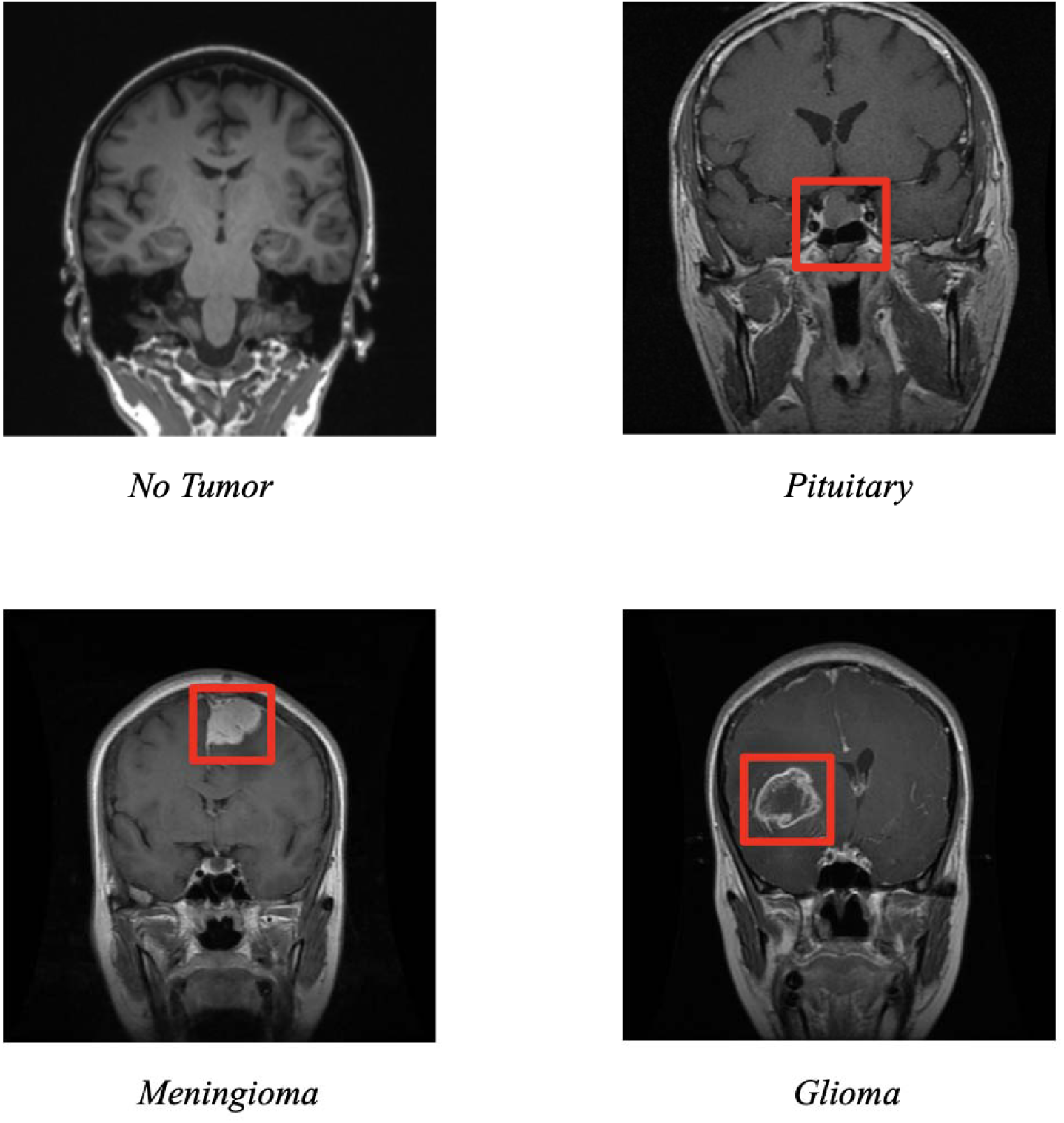
Images of the four different classes in the dataset, coronal view

## Methodology

The model that was implemented in this study was an Inception Resnet, a type of Convolutional Neural Network, or CNN, that has been trained on more than a million images from the ImageNet database and contains 164 layers^5^. CNNs are generally used to solve image classification problems such as the one at hand, and utilize a deep neural network to do so^6^. The structure of a CNN includes convolutional, pooling, and fully connected layers.

The convolutional layer utilizes small filters to scan through the given image and detect certain key features and patterns. It does this by comparing each n by n grid of pixels in the given image to the given filter’s pixel values. N represents the dimensions of the filter. The pixel values of the given filter are multiplied with the corresponding pixel values in the n by n grid on the image and are summed together to provide a value that represents the similarity between the filter and the area under inspection. This process is completed across all the n by n grids present in the image and with all the remaining filters.

Once this is done, the pooling layer downsamples the feature map given by the convolutional layer. This is done in order to more easily process the next convolutional layer. It works by dividing the output of the previous convolutional layer into smaller grids and then taking the maximum value from each grid to compose a smaller matrix.

Finally, the fully connected layer is responsible for predicting the final output while transforming the data into linear dimensions.

Before this Inception Resnet could be implemented, it was necessary to preprocess the data in order to minimize the effects of over and under fitting. More specifically, in order to minimize overfitting, a randomized training and testing split of the data was used. In addition to this, a random set of the data was either sheared, zoomed into, or flipped so that the final model would be able to accurately predict the class of the image even under unusual conditions. Once preprocessing was completed, the next step was to decide on the best hyperparameters to maximize validation metrics. This was done through a table of different combinations of parameters, listed below:

**Table 1:** Hyperparameters.

| A | B | C | D | E | F | G |
| --- | --- | --- | --- | --- | --- | --- |
|  | Dropout | Learning rate | # of steps | # of epochs | Training Acc | Testing Acc |
| 1 | 0.5 | 1.00E-05 | 150 | 30 | 86% | 73.30% |
| 2 | 0.25 | 1.00E-04 | 125 | 31 | 97.50% | 80% |
| 3 | 0.35 | 1.00E-06 | 170 | 32 | 53.30% | 33.30% |
| 4 | 0.15 | 1.00E-03 | 130 | 31 | 91.67% | 30% |
| 5 | 0.2 | 1.00E-04 | 120 | 31 | 97.50% | 73.30% |
| 6 | 0.3 | 1.00E-04 | 130 | 31 | 95.83% | 86.67% |
| 7 | 0.3 | 1.00E-04 | 130 | 60 | 100.00% | 80.00% |
| 8 | 0.3 | 1.00E-04 | 130 | 120 | 100.00% | 93.33% |
| 10 | 0.3 | 1.00E-04 | 130 | 150 | 99.17% | 96.67% |

When training the model, overfitting was prevented by splitting the dataset into a training and testing set. Data was split into 80% training and 20% testing respectively. These values were determined to be optimal through several tests. Through running the model with these different combinations, the last trial was found to yield the best testing accuracy, and thus the hyperparameters were finalized.

## Results and Discussion

Using the hyperparameters from the last trial, the testing accuracy was found to be 96.67%. This value was slightly less than the training accuracy, which was 99.17%. The training and testing losses were 0.019 and 0.0596, respectively.

Figure 3 shows an example of both a correctly and incorrectly classified image. In the example on the left, the model correctly predicts with 100% certainty that a pituitary tumor is present. However, on the right, the model is relatively unsure of its prediction of meningioma, and is ultimately wrong, as the image actually contains a glioma tumor. This is a good way to check the model on a case-by-case basis to see how it classifies individual images in the dataset. To further analyze the results of the model, loss graphs were produced.

**Figure 3:**
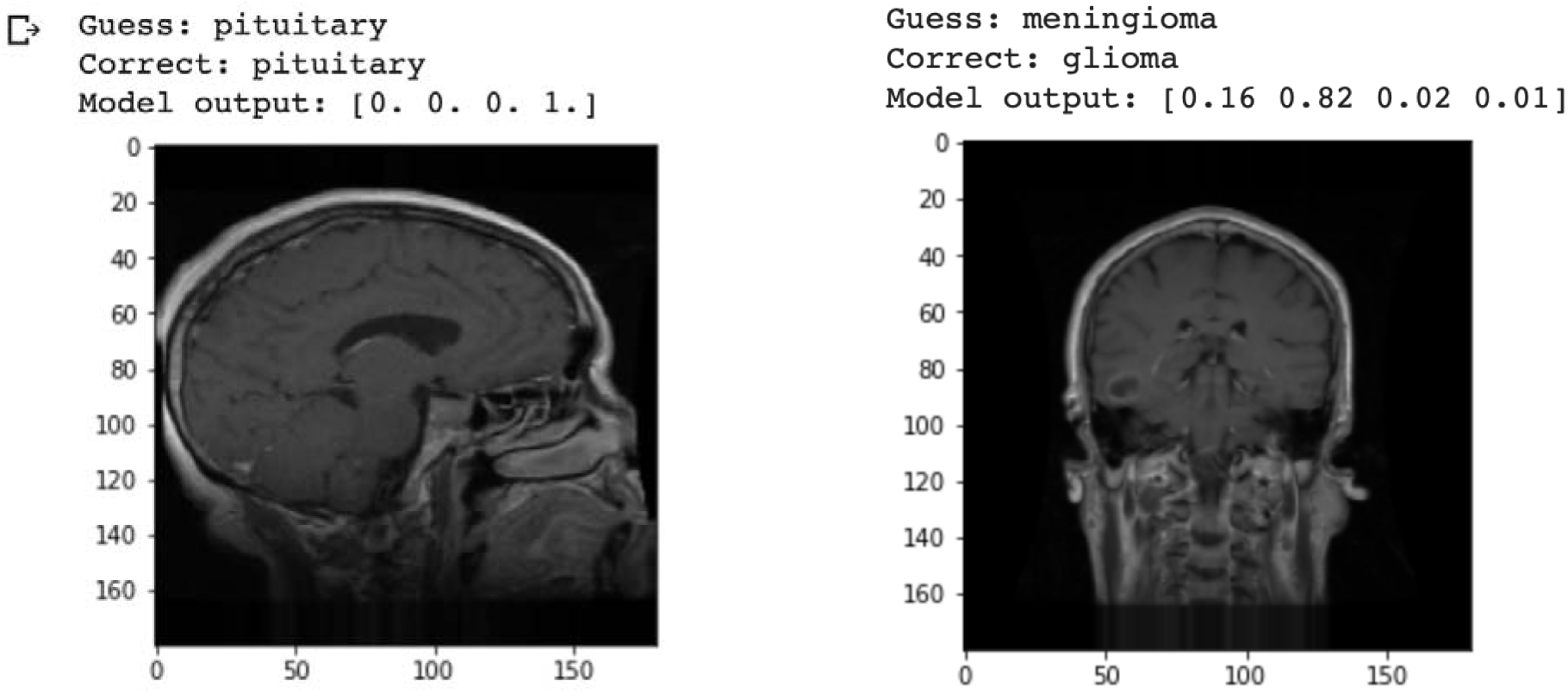
Model prediction of given images

The graph on the left in Figure 4 depicts the model’s loss curve going down exponentially as the number of epochs increases. Loss, in essence, represents a penalty for an incorrect prediction. There is a spike around epoch 100 as the model changes its approach to classifying the images, ultimately leading to a lower loss, meaning the model is quite accurate. This loss graph corresponds with the model’s accuracy graph shown on the right in Figure 4. As the loss curve decreases, the model’s accuracy increases, which makes sense as it becomes better at correctly classifying given images in the dataset.

**Figure 4:**
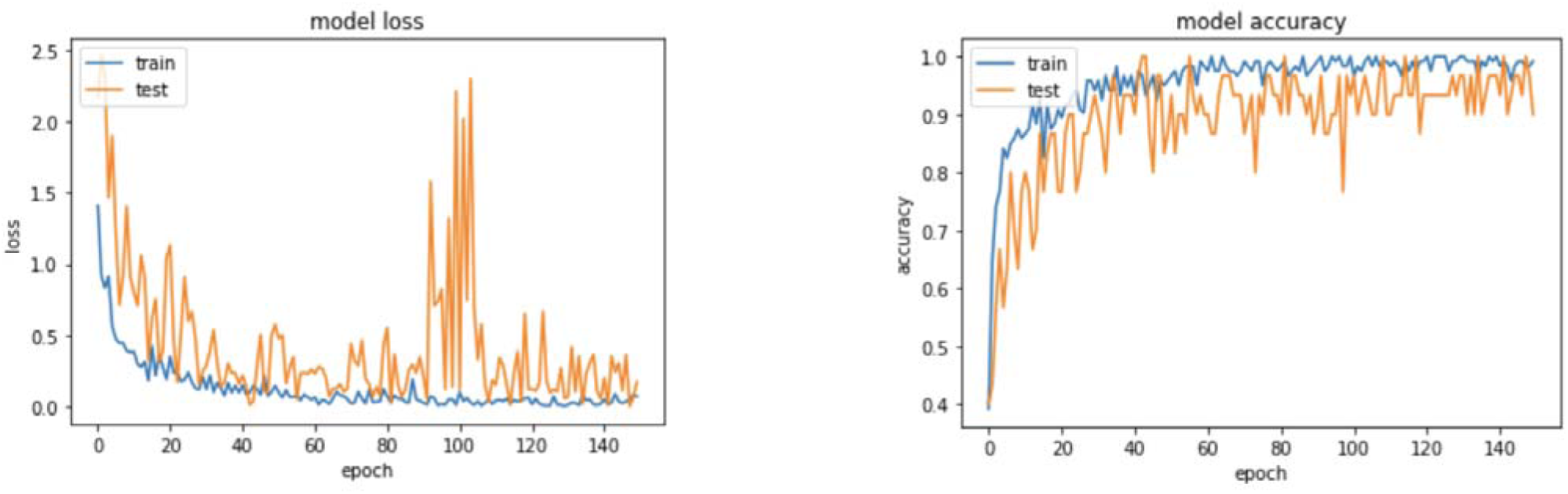
Model loss and accuracy graphs

While the accuracy measurement above is a good way of getting a broad overview of model performance, using a confusion matrix allows us to more closely inspect the results for each class. Thus, a confusion matrix was generated and is shown below in Figure 5, where each row represents the true label of the data, and each column represents the predicted label from the model output:

**Figure 5:**
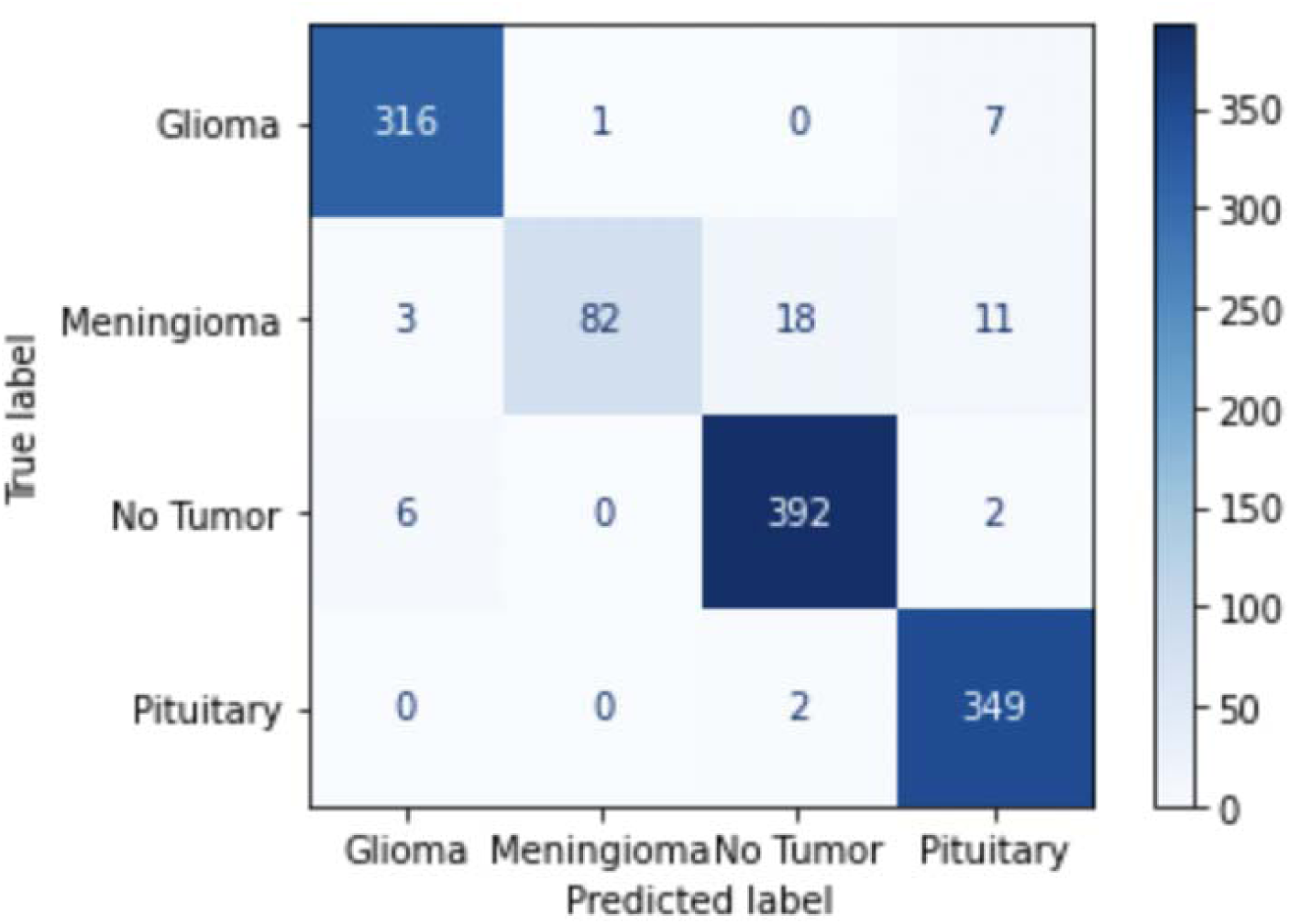
Confusion matrix of the trained Inception Resnet Model

**Table 2:** Sensitivities of tumors from confusion matrix.

| Tumor | Sensitivity |
| --- | --- |
| Glioma | 97.5% |
| Meningioma | 71.9% |
| No tumor | 98.0% |
| Pituitary | 99.4% |

Figure 5 is a visualization of the model’s performance which is called a confusion matrix. The matrix compares the model’s predicted labels of the tested image to their actual classes. Each row, if summed up, will result in the total number of tested images in that specific class. Based on this confusion matrix, it can be seen that the model performed best on determining if a given image either contained a glioma or pituitary tumor, or no tumor at all. However, in the meningioma class, the model only correctly predicted 82 out of 114 images, or 71.9% according to the sensitivity table, that contained a meningioma tumor. This low accuracy is most likely a result of the rarity of high grade meningioma tumors in the given dataset. There were only 114 meningioma images in the dataset compared to the large amounts of data points in the other classes, which explains this result.

It is also important to note that the model did not have many false negatives, which is beneficial in that it means there were not many cases where an image contained a tumor but was classified as having no tumor. More specifically, the model has a precision of 95% on images with no tumor(392/412), which indicates that only 5% of any kind of cancer cases were not detected. Other sources of possible error could come with the quality of the dataset and that there were some images that were not in the best quality or were taken at obscure angles.

## Conclusions

By training an Inception Resnet CNN model on the MRI scan dataset, I have created a brain tumor classification model that accurately predicts the presence and type of tumor in a brain MRI scan. This can now be implemented in healthcare settings.

In the future, it could be beneficial to incorporate other, more high quality datasets and test other forms of CNNs on the dataset to see if the accuracy can be reproduced and or improved.

## Data Availability

All data used in the study are available to the public online.

## Acknowledgments

I would like to acknowledge my mentor, Nic Thibodeaux, for helping me throughout the project in fixing various bugs and problems with the code.

## References

[1] “Key Statistics for Brain and Spinal Cord Tumors”(2023, January 12). American Cancer Society. https://www.cancer.org/cancer/brain-spinal-cord-tumors-adults/about/key-statistics.html

[2] Huang, J., Shlobin, N., Lam, S., DeCuypere, M., 2021, “Artificial Intelligence Applications in Pediatric Brain Tumor Imaging: A Systematic Review”, World Neurosurgery, pp. 99–105.https://www.sciencedirect.com/science/article/pii/S1878875021015655?scrlybrkr=e93f630c

[3] McFaline-Figueroa, J.R., Lee, E.Q., 2018, “Brain Tumors”, The American Journal of Medicine, pp. 874–882, https://www.sciencedirect.com/science/article/abs/pii/S0002934318300317

[4] Mambray, M.C., Barajas, R.F., Cha, S., 2015, “Modern Brain Tumor Imaging”, Synapse, pp. 8–23, https://synapse.koreamed.org/articles/1059671

[5] Szegedy, C., Ioffe, S., Vanhoucke, V., & Alemi, A. 2016, “Inception-v4, Inception-ResNet and the Impact of Residual Connections on Learning”. arXiv, 1602.07261. Retrieved from https://arxiv.org/abs/1602.07261v2

[6] Rawat, W., & Wang, Z. 2017, “Deep Convolutional Neural Networks for Image Classification: A Comprehensive Review”. Neural Comput., 29(9), 2352–2449. doi: 10.1162/neco_a_009

